# Factors Associated with the Outcomes of ST-Elevation Myocardial Infarction Patients in the ED

**DOI:** 10.1101/2022.10.30.22281696

**Authors:** Bill Hum, Yusef Shibly, Kamil Taneja, Karan Patel, Michael J. Diaz, Basil M. Baccouche, Tanisha Taneja, Sai Batchu, Aleem Mohamed, Alex Zhang, Hailey Hsiung, Urvish Patel

## Abstract

**Background:** ST-elevation myocardial infarction (STEMI) places a significant burden on the US healthcare system. However, there are gaps in our understanding of how patient demographics influence a STEMI’s risk to be admitted and the length of stay (LOS).

**Methods:** We conducted a retrospective analysis of the 2019 Nationwide Emergency Department Sample of patients with a primary diagnosis of STEMI. Multivariate regressions were used to determine factors associated with being admitted and longer length of stay (LOS).

**Results:** In 2019, 175,689 STEMI patients presented to the ED and 136,738 (77.8%) patients were admitted. Factors associated with higher risk of being admitted were coronary artery disease (OR:14.34, 95% confidence interval (CI): 12.43-16.54, p<0.001), modified Charlson Comorbidity Index (mCCI) of at least 3 (OR: 9.45, 95% CI: 7.33-12.17, p<0.001), and hyperlipidemia (OR:4.65, 95% CI:4.01-5.39, p<0.001). Black STEMI patients were less likely to be admitted than White STEMI patients (OR: 0.57, 95%CI: 0.43-0.75, p<0.001). Factors associated with a longer LOS include a mCCI of at least 3 (p<0.001), heart failure (p<0.001), and being an elderly patient (p<0.001). Black patients had a longer LOS than White patients (p<0.001). Medicaid beneficiaries were associated with a longer LOS than non-Medicaid beneficiaries (p<0.001).

**Conclusion:** Race and insurance status substantially affect a STEMI patient’s outcome in the ED.

## Introduction

An ST-elevation myocardial infarction (STEMI) is the complete thrombotic occlusion of the coronary arteries due to a ruptured atherosclerotic plaque with subsequent myocardial ischemia, injury, and/or necrosis. According to the 2022 update report by the American Heart Association, the estimated annual incidence of myocardial infarctions (MIs) is 605,000 new cases and 205,000 recurrent cases.^1^ Meanwhile, the prevalence of MIs between 2015 and 2018 in U.S. adults over the age of twenty was approximately 3.1%, or about 8.8 million people.^1^ Along with a high incidence, acute MIs were the 5th most expensive condition treated in U.S. hospitals in 2013, with an aggregate hospital cost of approximately $12.1 billion.^2^ MI is a significant burden on the US healthcare system, which warrants rigorous research on the outcomes of STEMI patients.

Racial disparity in medical outcomes and treatment that persists throughout the healthcare system. It is well known that after adjusting for socioeconomic status and severity of disease, there are major differences in the care provided to Blacks and non-White Hispanics when compared to non-Hispanic White patients.^3–5^ In the US, patient demographics (socioeconomic status (SES), race, and age) play a substantial role on a patient’s lifetime risk of a STEMI, treatment, and post-STEMI outcome.^6^ In particular, the Black and Hispanic patient groups are more likely to be given lower-quality MI care and suffer worse outcomes.^6^ For example, Black MI patients had longer door-to-drug, door-to-balloon, and transfer to revascularization hospital times compared to White MI patients.^7,8^

Even though there has been a significant amount of research done on the treatment care and quality of STEMI patients, there are still important gaps in the literature. To our knowledge, there are no research studies that have studied the relationship between patient demographics and hospital disposition (such as inpatient (IP) admission and length of stay (LOS)) in the context of STEMI. Given the high incidence and significant mortality of MIs, it is imperative to understand the patient factors that determine the outcome of their hospital stay, which can drastically impact their overall treatment.

In this study, we used a national ED database to study the relationship between patient and hospital characteristics that influence a STEMI patient’s outcomes, such as admission risk and LOS.

## Methods

The Nationwide Emergency Department Sample (NEDS) is the largest all-payer database for hospital-based emergency department (ED) visits. This database is supported and sponsored by the Healthcare Cost and Utilization Project (HCUP) and Agency for Healthcare Research and Quality. This database contains around 30 million patient records per year and contains roughly 20% of all U.S ED visits. The database is a large stratified cluster sample organized by hospital characteristics, such as region and teaching status. With linear regression models and sampling weights for each stratum, the NEDS can be used to extrapolate to the entire US ED population. Diagnoses in this database are listed using the International Classification of Diseases (ICD) coding system. The first ICD code is considered the primary diagnosis, for which all treatments are focused, and all other noted diagnoses are considered secondary diagnoses. More information about this dataset can be found on the HCUP website:https://www.hcup-us.ahrq.gov/nedsoverview.jsp.

We conducted a retrospective analysis on all patients with a STEMI (ICD-10: I20.0-I20.3) as the primary diagnosis in the 2019 NEDS database. Informed consent was not required because the dataset used de-identified patient information. All patient data was handed in according to the HCUP Data Use Agreement. Estimates less than or equal to 10 were not reported to protect patient confidentiality.

Linear regression and sampling weights of each stratum provided in the database were extrapolated from the NEDS database to the entire US ED population. Linear regression was used to estimate STEMI patient demographics (age, sex, race, median household income quartile based on patient’s zip code, insurance status), hospital characteristics (region, teaching status, trauma level), and associated comorbidities (obesity, hypertension, diabetes mellitus, hyperlipidemia, prior heart attack, prior stroke, coronary artery disease, chronic tobacco use, anti-platelet use, anti-thrombotic use, and anti-coagulant use). Patients were split into the following age groups: children (0-10), adolescents (11-20), young adults (21-44), adults (45-64), and the elderly (at least 65). Furthermore, multivariate logistic regression was used to calculate an adjusted odds ratio (OR) for the chance of being admitted while correcting for patient demographics, hospital characteristics, and comorbidities. To determine risk factors associated with longer LOS, a multivariate linear regression with the LOS as the dependent variable was used, while correcting for patient and hospital characteristics. A p-value less than 0.05 was considered significant.

## Results

In 2019, 175,689 patients presented to the ED with a primary diagnosis of STEMI. Although the sample population in this study was fairly heterogeneous, the majority of patients were white (74.1%), male (69.3%), paid with medicare insurance (44.8%), from the lowest income quartile (29.4%). Black (9.2%) and Hispanics (9.5%) comprised a percentage of patients. Most STEMI patients were Elderly (46.6%) or Adults (46.2%). A minority of patients were Young Adults (7.2%) or Adolescents (0.1%). In terms of hospital characteristics, most STEMI patients presented to Southern EDs (38.5%), metropolitan teaching hospitals (63.5%), and non-trauma centers (49.7%). The most common comorbidities in the patient population were coronary artery disease (66.9%), dyslipidemia (51.2%), tobacco use (47.2%), hypertension (43.7%), and diabetes mellitus (30.0%).A comprehensive list of patient demographics is available in Supplementary Table 1.

Figure 1 displays the outcomes of patients that presented to the ED (Figure 1A) and patients admitted to the hospital (Figure 1B). The most common outcomes from the ED were the IP setting (77.8%) and a discharge against medical advice (16.9%). The most common IP outcome was routine discharge (72.5%) and death (7.6%).

**Figure 1:**
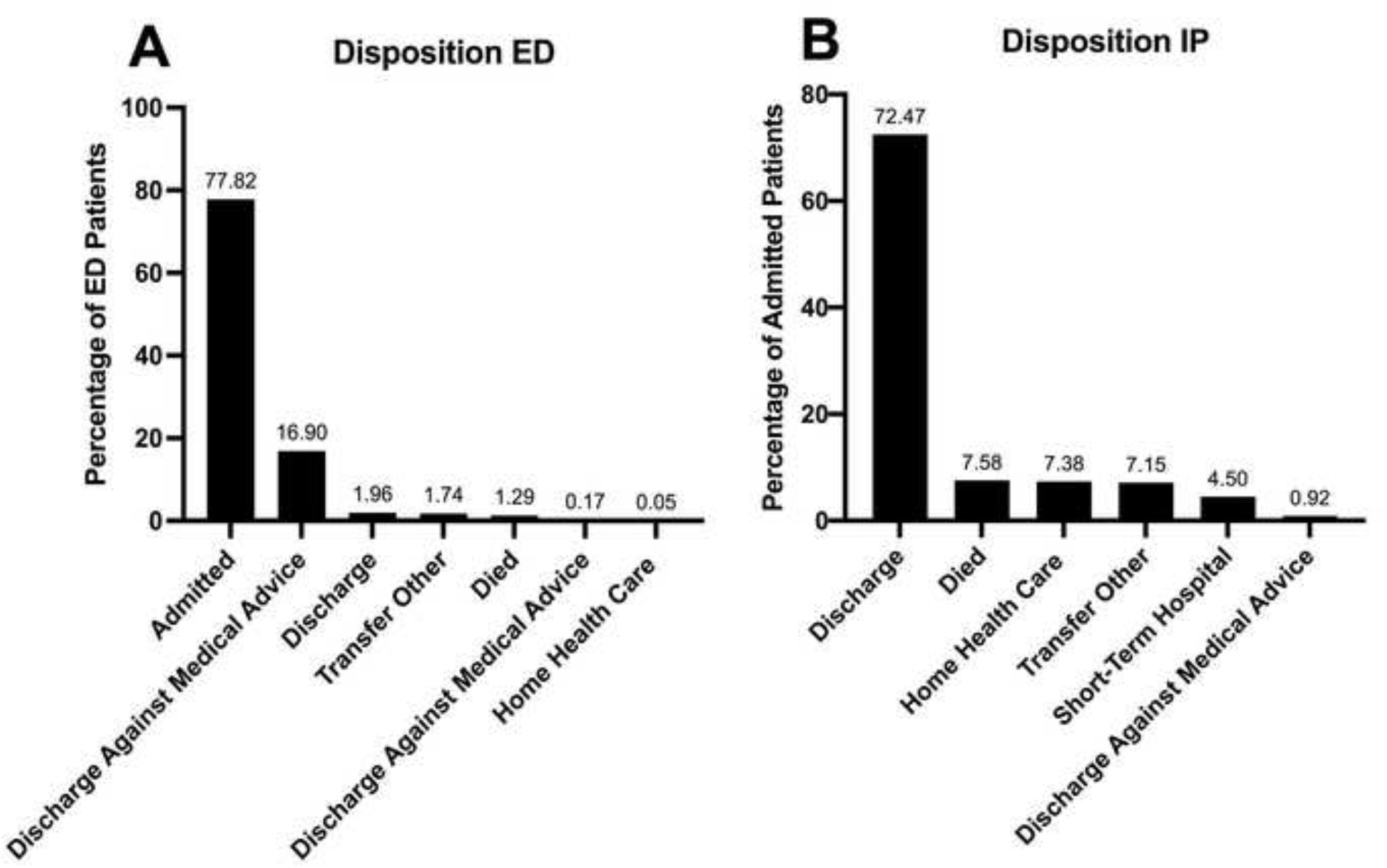
Disposition of STEMI Patients ED: Emergency Department IP: Inpatient Transfer Other: Includes Skilled Nursing Facility (SNF), Intermediate Care Facility (ICF), and another type of facility.

Out of the 175,689 patients that presented to the ED, 136,738 (77.8%) patients were admitted to the hospital. There were differences in odds of IP admittance within age, sex, race, hospital teaching, and hospital trauma levels. The highest risk factors (excluding patient comorbidities) for IP admission are displayed in Figure 2. Race played a substantial role in rates of admission. Black STEMI patients were less likely (OR: 0.57, 95% Confidence Interval (CI): 0.43-0.75, p<0.001) to be admitted than White patients. Patients that presented to a metropolitan teaching hospital were more likely to be admitted (OR: 2.04, 95% CI: 1.40-2.98, p<0.001) and patients that presented to a metropolitan were drastically less likely to be admitted (OR: 0.21, CI: 0.14-0.34, p<0.001) than a metropolitan non-teaching hospital. A comprehensive list of factors that affect a patient’s admission status is available in Supplementary Table 2.

**Figure 2:**
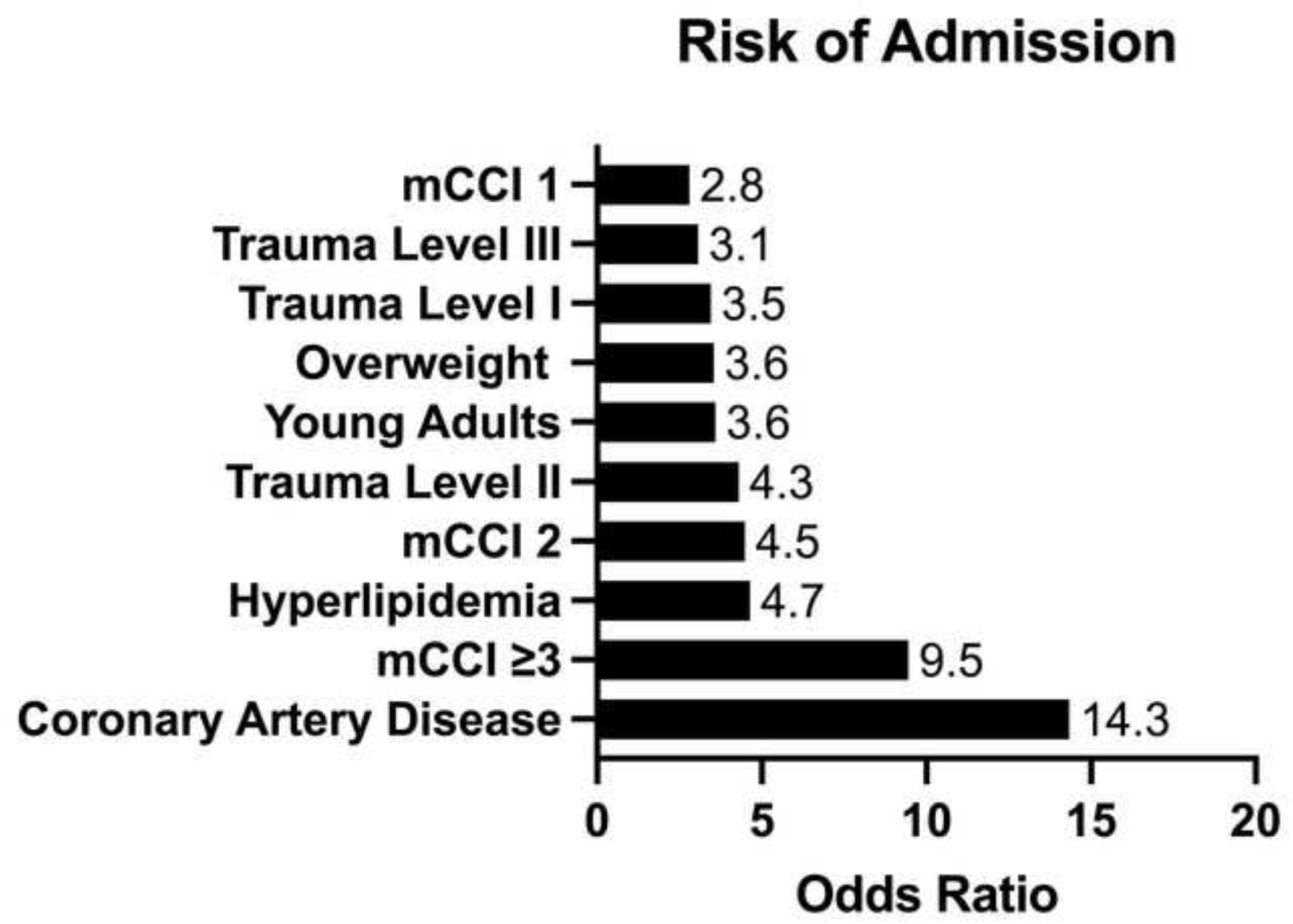
Factors Associated with Inpatient Admission Multivariate logistic regression was used to determine factors that are associated with higher admission risk. The regression included patient characteristics (e.g. comorbidities, demographics) and hospital characteristics (e.g. region, trauma status, and teaching status). The top 10 statistically significant risk factors are listed. mCCI: modified Charlson Comorbidity Index.

Among the admitted patients, the mean LOS was 3.8 ± 0.1 days. A patient’s LOS is highly related to patient factors (e.g. age, race, insurance status, income quartile) and hospital characteristics (teaching and trauma status. With the multivariate linear regression, White patients had a lower LOS than Black (p<0.001) and Asian/Pacific Islander (p=0.006) patients. White and Hispanic patients had the same LOS (p=0.052). Furthermore, patients with Medicaid had a longer LOS than Medicare patients (p<0.001). There was no difference between Medicare and other payment methods (e.g. private insurance and uninsured). The factors (outside of patient comorbidities) that most significantly prolong a patient’s stay are displayed in Figure 3. A comprehensive list of factors that affect a patient’s LOS and mean LOS per group is available in Supplementary Table 3.

**Figure 3:**
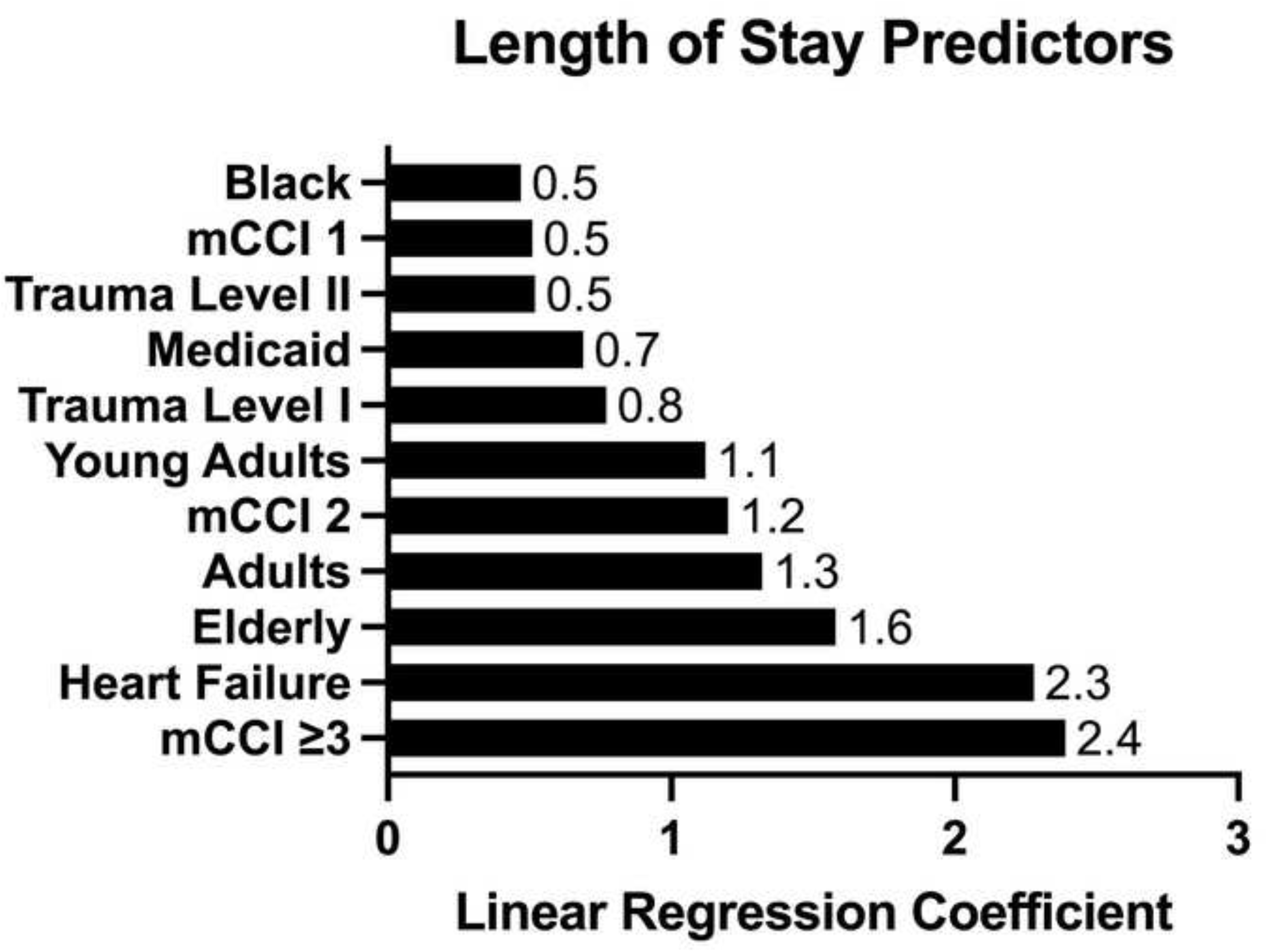
Factors Associated with Longer Hospital Stays Multivariate linear regression was used to determine factors associated with longer hospital stays. The regression included patient characteristics (e.g. comorbidities, demographics) and hospital characteristics (e.g. region, trauma status, and teaching status). The top 10 statistically significant risk factors are listed. mCCI: modified Charlson Comorbidity Index.

## Discussion

In this study, we used a national ED database to analyze the outcomes of STEMI patients in the ED. We demonstrated that a patient’s race is intimately related to LOS and risk of IP admittance. Furthermore, insurance status plays a substantial role in a patient’s LOS. This study adds to the rapidly growing literature that highlights how patient demographics determine the quality of care in the US healthcare system.^3–5^

In our study, we showed that payer status was predictive of a patient’s LOS. Specifically, we showed that Medicaid status was associated with the highest LOS when compared to Medicaid and other payment methods. This is very alarming since Medicaid is an important source of financial aid for low income, non-elderly patients. According to the Center of Medicare and Medicaid Services, over 70 million individuals were enrolled in Medicaid in 2019.^9^ However, literature in the past few years is indicating that Medicaid status is associated with lower quality care. ^10–12^. It is important to note that the association has It is possible that Medicaid patients present to the ED in worse conditions, have weak social support, racial differences, mistrust of the medical system, and cultural/language barriers.^6,13–17^ It is important to note that Medicaid insurance status is associated with longer LOS and we have not proven a causal link between long LOS and Medicaid status. We believe that the aforementioned factors can help explain why Medicaid beneficiaries have longer LOS. Medicaid’s expansion was important in improving healthcare access to low-income individuals and there is evidence suggesting that the expansion has decreased cardiovascular mortality.^18,19^ However, as our findings suggest, there is considerable amount of work that needs to be done to truly minimize the outcome disparity between Medicaid and non-Medicaid patients.

Furthermore, our data reinforce that finding that race determines a patient’s outcome. In our study, we demonstrated that Black STEMI patients were 43% less likely to be admitted than White STEMI patients and had longer LOS than White patients, even after correcting for demographics, income, comorbidities, insurance status, and hospital characteristics. Along with lower admission rates, Black MI patients have lower revascularization rates, rate of transfer to hospitals with revascularization suites, and door-to-balloon times.^6–8^Along with differences of care, it is possible that Black patients present to the ED with more severe conditions than White patients. While we can correct for the presence of a comorbidity, we are not able to correct for the initial STEMI severity. It is well known that metabolic syndrome (and its associated conditions) and cardiovascular events are more common in Black patient.^6,20–22^. Worse initial STEMI severity along with lower quality of care of Black patients can also explain the longer LOS of Black STEMI patients.

This study’s strengths include its large size, access to hospital characteristics, patient demographics, and recency of data. However, this study has a few important issues. The NEDS is an encounter-level database which does not capture patients that are transferred from an ED from one hospital to the IP setting to another hospital. It will only track a patient that was admitted in the same hospital. Furthermore, the database does not provide granular clinical information (such as cardiac marker levels, lab values) which would help us characterize the initial severity of the patient. The database does not provide data on complications of care, which would ideally be included in our multivariate analysis for LOS. Also, the NEDS does not use updated guidelines on racial and ethnic terminology, which can its conclusion difficult to understand. ^23^

## Conclusion

STEMI places a substantial burden on the US healthcare system. Throughout the medical system, a large amount of evidence indicates that race, ethnicity, socioeconomic status, and insurance status greatly influence the health of an individual and treatment outcomes. In this study, we add upon the literature field. We demonstrated that race impacts both the odds of being admitted to the hospital for a STEMI and LOS. Specifically, Black patients are 43% less likely to be admitted to the hospital when compared to White Patients, after correcting for numerous factors. Furthermore, insurance status, an indicator of socioeconomic status, is related to the LOS of a patient. We found that Medicaid beneficiaries had longer LOS than non-Medicaid beneficiaries. Future studies are called upon to investigate the long-term progress of STEMI patients across insurance and racial groups to fully evaluate the impact of the disparities. We hope that our findings can be used to change health care policies to minimize the disparities and provide equal care to all patients.

## Data Availability

Available for purchase at the following link: https://www.hcup-us.ahrq.gov/nedsoverview.jsp

## Figure Legends

**Supplementary Table 1:** Demographics of Patients with a Primary ST-Elevation Myocardial Infarction

| Characteristic | Adolescent<br>s N<sup>a</sup> (%) | Young<br>Adults<br>N<sup>a</sup> (%) | Adults<br>N<sup>a</sup> (%) | Elderly<br>N<sup>a</sup> (%) | Total<br>N<sup>a</sup> (%) |
| --- | --- | --- | --- | --- | --- |
| **Sex** |  |  |  |  |  |
| Male | 104 (74.2) | 9936 (78.7) | 61776 (76.1) | 49888 (61) | 121704 (69.3) |
| Female | 36 (25) | 2688 (21.3) | 19363 (23.9) | 31898 (39) | 53985 (30.7) |
| **Race** |  |  |  |  |  |
| White | 61 (45.6) | 7605 (61.8) | 56388 (71.3) | 63123 (78.8) | 127177 (74.1) |
| Black | 20 (14.9) | 2108 (17.1) | 8104 (10.2) | 5645 (7) | 15877 (9.2) |
| Hispanic | 48 (36.2) | 1506 (12.2) | 8561 (10.8) | 6214 (7.8) | 16330 (9.5) |
| Asian or Pacific Islander | <sup>-b</sup> | 510 (4.1) | 2441 (3.1) | 2198 (2.7) | 5149 (3.0) |
| Native American | <sup>-b</sup> | 93 (0.8) | 387 (0.5) | 219 (0.3) | 699 (0.4) |
| Other | <sup>-b</sup> | 491 (4) | 3216 (4.1) | 2746 (3.4) | 6457 (3.8) |
| **Primary Payer** |  |  |  |  |  |
| Medicare | <sup>-b</sup> | 676 (5.4) | 9268 (11.4) | 68699 (84) | 78644 (44.8) |
| Medicaid | 50 (35.9) | 3010 (23.9) | 13048 (16.1) | 1191 (1.5) | 17298 (9.9) |
| Private Insurance | 67 (47.8) | 6024 (47.8) | 43921 (54.2) | 9097 (11.1) | 59109 (33.7) |
| Self-Pay | 23 (16.3) | 2289 (18.2) | 10597 (13.1) | 1218 (1.5) | 14126 (8) |
| No Charge | <sup>-b</sup> | 175 (1.4) | 899 (1.1) | 77 (0.1) | 1151 (0.7) |
| Other | <sup>-b</sup> | 427 (3.4) | 3259 (4) | 1476 (1.8) | 5161 (2.9) |
| **Quartile of Median Household**<br>**Income of Zip Code** |  |  |  |  |  |
| First | 32 (24.1) | 4137 (33.4) | 24429 (30.7) | 21970 (27.4) | 50568 (29.4) |
| Second | 29 (21.8) | 3297 (26.6) | 20198 (25.4) | 20816 (26) | 44340 (25.7) |
| Third | 28 (21.5) | 2947 (23.8) | 18464 (23.2) | 19730 (24.6) | 41169 (23.9) |
| Fourth | 43 (32.5) | 2021 (16.3) | 16471 (20.7) | 17631 (22) | 36165 (21) |
| **Hospital Region** |  |  |  |  |  |
| Northeast | 21 (15.1) | 1978 (15.7) | 14089 (17.4) | 14923 (18.2) | 31012 (17.7) |
| Midwest | 23 (16.1) | 3228 (25.6) | 19510 (24) | 19301 (23.6) | 42061 (23.9) |
| South | 60 (42.5) | 5194 (41.1) | 31851 (39.3) | 30615 (37.4) | 67720 (38.5) |
| West | 37 (26.3) | 2224 (17.6) | 15694 (19.3) | 16947 (20.7) | 34902 (19.9) |
| **Hospital Teaching Status** |  |  |  |  |  |
| Metropolitan Non-teaching | 26 (18.5) | 2720 (21.5) | 16863 (20.8) | 16463 (20.1) | 36072 (20.5) |
| Metropolitan Teaching | 105 (74.7) | 7905 (62.6) | 51964 (64) | 51618 (63.1) | 111592 (63.5) |
| Non-metropolitan | <sup>-b</sup> | 1999 (15.8) | 12318 (15.2) | 13704 (16.8) | 28031 (16) |

| **Hospital Trauma Level Designation** |  |  |  |  |  |
| --- | --- | --- | --- | --- | --- |
| Non-Trauma | 73 (52.2) | 6399 (50.9) | 39777 (49.2) | 40798 (50.1) | 87047 (49.7) |
| Level I | 14 (10.1) | 1947 (15.5) | 13266 (16.4) | 12005 (14.7) | 27232 (15.6) |
| Level II | 31 (22) | 2165 (17.2) | 15533 (19.2) | 15791 (19.4) | 33521 (19.1) |
| Level III | 22 (15.8) | 2004 (15.9) | 12028 (14.9) | 12573 (15.4) | 26627 (15.2) |
| **Co-Morbidities** |  |  |  |  |  |
| Overweight | 15 (10.8) | 2972 (23.5) | 14802 (18.2) | 8921 (10.9) | 26711 (15.2) |
| Hypertensive | -<sup>b</sup> | 5020 (39.8) | 37365 (46) | 34401 (42.1) | 76795 (43.7) |
| Diabetes | -<sup>b</sup> | 3004 (23.8) | 23867 (29.4) | 25888 (31.7) | 52768 (30) |
| Hyperlipidemia | 21 (14.7) | 5131 (40.6) | 41351 (51) | 43440 (53.1) | 89942 (51.2) |
| Old Heart Attack | -<sup>b</sup> | 1143 (9.1) | 9296 (11.5) | 9310 (11.4) | 19753 (11.2) |
| Heart Failure | -<sup>b</sup> | 1746 (13.8) | 14554 (17.9) | 21891 (26.8) | 38198 (21.7) |
| Old Stroke | -<sup>b</sup> | 230 (1.8) | 2776 (3.4) | 5679 (6.9) | 8684 (4.9) |
| Coronary | 19 (13.3) | 7510 (59.5) | 54620 (67.3) | 55449 (67.8) | 117598 (66.9) |
| Tobacco | 23 (16.6) | 7358 (58.3) | 43342 (53.4) | 32139 (39.3) | 82863 (47.2) |
| Erectile Dysfunction | -<sup>b</sup> | 24 (0.2) | 339 (0.4) | 241 (0.3) | 604 (0.3) |
| Antiplatelets/<br>Antithrombotics | -<sup>b</sup> | 786 (6.2) | 6091 (7.5) | 7059 (8.6) | 13936 (7.9) |
| Anticoagulants | -<sup>b</sup> | 320 (2.5) | 2600 (3.2) | 5527 (6.8) | 8452 (4.8) |
| Thrombolytics | -<sup>b</sup> | 313 (2.5) | 1409 (1.7) | 1276 (1.6) | 2998 (1.7) |
| **Modified Charlson Co-Morbidity Index** |  |  |  |  |  |
| 0 | 126 (90) | 6675 (52.9) | 36183 (44.6) | 27437 (33.5) | 70421 (40.1) |
| 1 | -<sup>b</sup> | 3838 (30.4) | 25175 (31) | 22241 (27.2) | 51258 (29.2) |
| 2 | -<sup>b</sup> | 1226 (9.7) | 10178 (12.5) | 12602 (15.4) | 24009 (13.7) |
| 3 | -<sup>b</sup> | 885 (7) | 9609 (11.8) | 19506 (23.8) | 30007 (17.1) |

<sup>a</sup>Number of patients
<sup>b</sup>As per Health Care and Utilization Project Data Use Agreements, estimates less than or equal to 10 not reported.

**Supplementary Table 2:** Risk of Inpatient Admission

| Characteristic | N<sup>a</sup> (%) | OR<sup>b</sup> (95% CI<sup>c</sup>) | p |
| --- | --- | --- | --- |
| **Age** |  |  |  |
| Adolescents | 42 (0) | Reference |  |
| Young Adults | 9466 (6.9) | 3.6 (1.09-11.75) | 0.036 |
| Adults | 63545 (46.5) | 2.9 (0.88-9.58) | 0.082 |
| Elderly | 63685 (46.6) | 2.2 (0.66-7.37) | 0.199 |
| **Gender** |  |  |  |
| Male | 94832 (69.4) | Reference |  |
| Female | 41901 (30.6) | 1.19 (1.09-1.3) | <0.001 |
| **Race** |  |  |  |
| White | 97878 (73.2) | Reference |  |
| Black | 12227 (9.1) | 0.57 (0.43-0.75) | <0.001 |
| Hispanic | 13073 (9.8) | 0.79 (0.61-1.03) | 0.085 |
| Asian or Pacific Islander | 4413 (3.3) | 1.05 (0.76-1.47) | 0.758 |
| Native American | 556 (0.4) | 1.17 (0.52-2.64) | 0.7 |
| Other | 5648 (4.2) | 1.58 (1.1-2.27) | 0.013 |
| **Primary Payer** |  |  |  |
| Medicare | 61466 (45) | Reference |  |
| Medicaid | 13584 (9.9) | 1.09 (0.9-1.32) | 0.388 |
| Private Insurance | 46365 (33.9) | 1.04 (0.86-1.25) | 0.712 |
| Self-Pay | 10231 (7.5) | 0.82 (0.64-1.06) | 0.129 |
| No Charge | 1022 (0.7) | 2.4 (1.25-4.62) | 0.008 |
| Other | 3951 (2.9) | 1.07 (0.8-1.41) | 0.659 |
| **Quartile of Median Household Income of Zip code** |  |  |  |
| First | 36839 (27.5) | Reference |  |
| Second | 33255 (24.8) | 0.95 (0.76-1.19) | 0.682 |
| Third | 32715 (24.4) | 0.84 (0.64-1.1) | 0.203 |
| Fourth | 31293 (23.3) | 1.12 (0.82-1.53) | 0.489 |
| **Hospital Region** |  |  |  |
| Northeast | 24985 (18.3) | Reference |  |
| Midwest | 30766 (22.5) | 0.74 (0.47-1.16) | 0.187 |
| South | 53563 (39.2) | 1.43 (0.87-2.33) | 0.154 |
| West | 27424 (20.1) | 0.8 (0.48-1.32) | 0.382 |
| **Hospital Teaching Status** |  |  |  |
| Metropolitan Non-teaching | 26360 (19.3) | Reference |  |
| Metropolitan Teaching | 100079 (73.2) | 2.04 (1.4-2.98) | <0.001 |
| Non-metropolitan | 10299 (7.5) | 0.21 (0.14-0.34) | <0.001 |
| **Hospital Trauma Level Designation** |  |  |  |
| Non-Trauma | 56114 (41.2) | Reference |  |
| Level I | 25614 (18.8) | 3.45 (1.5-7.94) | 0.004 |
| Level II | 31613 (23.2) | 4.3 (2.65-6.96) | <0.001 |
| Level III | 22191 (16.3) | 3.07 (2.04-4.62) | <0.001 |
| **Co-Morbidities** |  |  |  |
| Coronary Artery Disease | 111738 (81.7) | 14.34 (12.43-16.54) | <0.001 |
| Hyperlipidemia | 84498 (61.8) | 4.65 (4.01-5.39) | <0.001 |
| Overweight | 25363 (18.5) | 3.55 (2.91-4.32) | <0.001 |
| Heart Failure | 36173 (26.5) | 2.23 (1.87-2.66) | <0.001 |
| Thrombolytics | 2298 (1.7) | 1.91 (1.15-3.17) | 0.012 |
| Antiplatelets/Antithrombotics | 13058 (9.5) | 1.58 (1.2-2.08) | 0.001 |
| Tobacco | 69106 (50.5) | 1.4 (1.24-1.58) | <0.001 |
| Hypertension | 61662 (45.1) | 1.3 (1.16-1.45) | <0.001 |
| Old Stroke | 7457 (5.5) | 1.07 (0.86-1.33) | 0.528 |
| Anticoagulants | 6971 (5.1) | 0.77 (0.62-0.94) | 0.012 |
| Erectile Dysfunction | 552 (0.4) | 0.74 (0.35-1.56) | 0.427 |
| Old Heart Attack | 17054 (12.5) | 0.43 (0.36-0.51) | <0.001 |
| Diabetes | 44921 (32.9) | 0.33 (0.29-0.38) | <0.001 |
| **Modified Charlson Co-Morbidity Index** |  |  |  |
| 0 | 43878 (32.1) | Reference |  |
| 1 | 42753 (31.3) | 2.8 (2.41-3.26) | <0.001 |
| 2 | 21833 (16) | 4.49 (3.65-5.53) | <0.001 |
| 3 | 28274 (20.7) | 9.45 (7.33-12.17) | <0.001 |

<sup>a</sup>Number of patients
<sup>b</sup>Odds ratio
<sup>c</sup>Confidence interval

**Supplementary Table 3:** Factors Associated with Hospital Length of Stay

| Characteristic | Mean (SE<sup>a</sup>) | $\beta^b$ (95% CI<sup>c</sup>) | p |
| --- | --- | --- | --- |
| **Age** |  |  |  |
| Adolescents | 2.4 (0.6) | Reference |  |
| Young Adults | 3.3 (0.1) | 1.12 (0.25-2) | 0.012 |
| Adults | 3.6 (0.1) | 1.32 (0.46-2.18) | 0.003 |
| Elderly | 4.2 (0.1) | 1.58 (0.7-2.47) | <0.001 |
| **Gender** |  |  |  |
| Male | 3.8 (0.1) | Reference |  |
| Female | 4 (0.1) | -0.1 (-0.23-0.02) | 0.105 |
| **Race** |  |  |  |
| White | 3.7 (0.0) | Reference |  |
| Black | 4.5 (0.2) | 0.47 (0.2-0.75) | 0.001 |
| Hispanic | 4.1 (0.1) | 0.23 (0-0.46) | 0.052 |
| Asian or Pacific Islander | 4.2 (0.1) | 0.36 (0.07-0.66) | 0.014 |
| Native American | 4.1 (0.4) | 0.52 (-0.24-1.28) | 0.182 |
| Other | 4.4 (0.2) | 0.48 (0.13-0.83) | 0.007 |
| **Primary Payer** |  |  |  |
| Medicare | 4.1 (0.1) | Reference |  |
| Medicaid | 4.3 (0.1) | 0.69 (0.42-0.96) | <0.001 |
| Private Insurance | 3.4 (0.1) | 0.18 (-0.04-0.39) | 0.114 |
| Self-Pay | 3.4 (0.1) | 0.13 (-0.16-0.41) | 0.386 |
| No Charge | 3.7 (0.5) | 0.44 (-0.46-1.34) | 0.338 |
| Other | 3.8 (0.2) | 0.28 (-0.05-0.61) | 0.100 |
| **Quartile of Median Household Income of Zip code** |  |  |  |
| First | 4 (0.1) | Reference |  |
| Second | 3.7 (0.1) | -0.2 (-0.37--0.03) | 0.022 |
| Third | 3.8 (0.1) | -0.16 (-0.33-0.01) | 0.064 |
| Fourth | 3.8 (0.1) | -0.18 (-0.39-0.03) | 0.098 |
| **Hospital Region** |  |  |  |
| Northeast | 4.3 (0.1) | Reference |  |
| Midwest | 3.7 (0.1) | -0.35 (-0.61--0.09) | 0.009 |
| South | 3.8 (0.1) | -0.18 (-0.46-0.1) | 0.214 |
| West | 3.7 (0.1) | -0.57 (-0.88--0.26) | <0.001 |
| **Hospital Teaching Status** |  |  |  |
| Metropolitan Non-teaching | 3.4 (0.1) | Reference |  |
| Metropolitan Teaching | 4 (0.1) | 0.14 (-0.07-0.35) | 0.198 |
| Non-metropolitan | 3.3 (0.2) | -0.26 (-0.57-0.05) | 0.096 |
| **Hospital Trauma Level Designation** |  |  |  |
| Non-Trauma | 3.6 (0.1) | Reference |  |
| Level I | 4.6 (0.1) | 0.77 (0.48-1.06) | <0.001 |
| Level II | 4 (0.1) | 0.52 (0.29-0.74) | <0.001 |
| Level III | 3.5 (0.1) | 0.11 (-0.13-0.35) | 0.382 |
| **Co-Morbidities** |  |  |  |
| Overweight | 3.9 (0.1) | 0.13 (-0.02-0.28) | 0.082 |
| Hypertension | 2.9 (0) | -0.02 (-0.11-0.08) | 0.726 |
| Diabetes | 4.4 (0.1) | -0.35 (-0.52--0.18) | <0.001 |
| Hyperlipidemia | 3.6 (0) | -0.52 (-0.65--0.38) | <0.001 |
| Old Heart Attack | 3.7 (0.1) | -0.38 (-0.53--0.22) | <0.001 |
| Heart Failure | 6.2 (0.1) | 2.28 (2.06-2.5) | <0.001 |
| Old Stroke | 4 (0.1) | -0.35 (-0.55--0.15) | 0.001 |
| Coronary | 3.8 (0.1) | -0.004 (-0.22-0.22) | 0.971 |
| Tobacco | 3.5 (0) | -0.61 (-0.73--0.49) | <0.001 |
| Erectile Dysfunction | 3.3 (0.2) | -0.32 (-0.82-0.18) | 0.209 |
| Antiplatelets/Antithrombotics | 3.7 (0.1) | -0.36 (-0.52--0.21) | <0.001 |
| Anticoagulants | 4.3 (0.1) | -0.12 (-0.36-0.13) | 0.344 |
| Thrombolytics | 4.1 (0.2) | 0.29 (-0.08-0.65) | 0.127 |
| **Modified Charlson Co-Morbidity Index** |  |  |  |
| 0 | 2.5 (0) | Reference |  |
| 1 | 3.5 (0.1) | 0.51 (0.38-0.63) | <0.001 |
| 2 | 4.5 (0.1) | 1.2 (1-1.39) | <0.001 |
| 3 | 5.9 (0.1) | 2.39 (2.14-2.64) | <0.001 |

<sup>a</sup>Standard error
<sup>b</sup>Linear Regression Coefficient
<sup>c</sup>Confidence interval
<sup>d</sup>As per Health Care and Utilization Project Data Use Agreements, estimates less than or equal to 10 not reported

## Declaration of interests

✉The authors declare that they have no known competing financial interests or personal relationships that could have appeared to influence the work reported in this paper.

□The authors declare the following financial interests/personal relationships which may be considered as potential competing interests:

## Notes

### Competing Interest Statement

The authors have declared no competing interest.

### Funding Statement

This study did not receive any funding

### Author Declarations

The data used used de-identified data. Therefore, IRB approval was not required. The owner of the data (The Healthcare Utilization Project) gave ethical approval for this work.

